# Interpretable Brain Disease Classification and Relevance-Guided Deep Learning

**DOI:** 10.1101/2021.09.09.21263013

**Authors:** Christian Tinauer, Stefan Heber, Lukas Pirpamer, Anna Damulina, Reinhold Schmidt, Rudolf Stollberger, Stefan Ropele, Christian Langkammer

**Author notes:** Correspondence, Department of Neurology, Medical University of Graz, Auenbruggerplatz 22, 8036 Graz, Austria.

## Abstract

Deep neural networks are increasingly used for neurological disease classification by MRI, but the networks’ decisions are not easily interpretable by humans. Heat mapping by deep Taylor decomposition revealed that (potentially misleading) image features even outside of the brain tissue are crucial for the classifier’s decision. We propose a regularization technique to train convolutional neural network (CNN) classifiers utilizing relevance-guided heat maps calculated online during training. The method was applied using T1-weighted MR images from 128 subjects with Alzheimer’s disease (mean age=71.9±8.5 years) and 290 control subjects (mean age=71.3±6.4 years). The developed relevance-guided framework achieves higher classification accuracies than conventional CNNs but more importantly, it relies on less but more relevant and physiological plausible voxels within brain tissue. Additionally, preprocessing effects from skull stripping and registration are mitigated. With the interpretability of the decision mechanisms underlying CNNs, these results challenge the notion that unprocessed T1-weighted brain MR images in standard CNNs yield higher classification accuracy in Alzheimer’s disease than solely atrophy.

## Introduction

Alzheimer’s disease (AD) is the most common form of dementia with about 50 million patients and a substantial burden for our healthcare systems, caregivers and next of kin (Scheltens et al., 2021). While postmortem diagnosis can be obtained from the histological examination of tissue samples from affected anatomical regions (Braak et al., 2006; Braak and Braak, 1991), in vivo diagnosis is hampered by clinical symptom similarities and its accuracy is rather low (71%–87% sensitivity and 44%–71% specificity) (Oldan et al., 2021).

In addition to clinical and neuropsychological tests, medical imaging is increasingly used to strengthen diagnosis by PET imaging ligands to amyloid-β and tau proteins combined with MRI. Recently revised diagnosis criteria for AD are clinical-biological and require both clinical phenotype and biomarker evidence (Aβ or tau) of AD (Dubois et al., 2021). Although the presence of extracellular neuritic Aβ plaques is part of several diagnosis criteria their clinical value is discussed controversially, whereas selective tau ligands do reflect clinical severity and memory impairment and also serve for invivo Braak-staging (Biel et al., 2021). Based on imaging tau pathology, recent fascinating data-driven work found that tau-PET can be used to identify four spatiotemporal phenotypes which exhibit different clinical profiles and longitudinal outcomes and thus opens an avenue for personalized treatment (Vogel et al., 2021). However, AD has a long prodromal and asymptomatic inflammatory phase where radioactive PET tracers cannot be used as a means for its prognosis in a healthy population. Because pathological changes are occurring decades before initial clinical manifestations, early biomarkers in a broad population might be obtained best by MRI, where volumetry and especially hippocampal atrophy are presently used as imaging markers (Henneman et al., 2009; Leung et al., 2013; Sluimer et al., 2008).

Deep learning is omnipresent in medical imaging, including image reconstruction (Hammernik et al., 2018), segmentation (Kleesiek et al., 2016), and classification (Esteva et al., 2017; Bäckström et al., 2018). Convolutional neural networks (CNNs) are utilized for neurological disease classification (Noor et al., 2020; Vieira et al., 2017; Zhang et al., 2020) and regression (Dinsdale et al., 2021a) in prevalent neurological disorders such as Alzheimer’s disease (Oh et al., 2019; Bäckström et al., 2018; Böhle et al., 2019; Korolev et al., 2017), Parkinson’s disease (Karapinar Senturk, 2020) and multiple sclerosis (Eitel et al., 2019).

Despite their improved performance, those models are generally not easily interpretable by humans and deep neural networks (DNNs) are mostly seen as black boxes where data in combination with extensive learning efforts yields decisions (Davatzikos, 2019). One striking example of misguided feature extraction of DNNs is described in (Lapuschkin et al., 2019), where secondary photo watermarks identified horses better than the actual animal print. In the context of brain MRI it has been shown that learned features for age estimation are influenced by the applied registration type (linear vs. nonlinear) (Dinsdale et al., 2021a). However, no systematic investigation of the preprocessing of brain MR images for disease classification with CNNs has been conducted, but the studies (Böhle et al., 2019; Eitel et al., 2019; Oh et al., 2019) aimed at explaining their applied classifier. Preprocessing is a crucial step, with skull stripping (brain extraction) creating artificial edges and interpolation and regridding necessary for registration. CNNs can incorporate these newly introduced features during training and base their classification results thereon.

Medical imaging has high legal requirements as e.g. the EU’s General Data Protection Regulation (GDPR) explicitly requires the right to explanation for users subjected to decisions of an automated processing system (Goodman and Flaxman, 2017) and the US are endorsing the OECD AI Principles of transparency and explainability (OECD, 2019). Consequently, medical decision-supporting algorithms require verifying that this is not the result of exploiting data artifacts and that the high accuracy of classification decisions are explainable to avoid biased results (Lapuschkin et al., 2019, 2016). In the present work we used heat (or saliency) mapping, which is enabling perceptive interpretability to explain a classification result in terms of maps overlaid on the input (Tjoa and Guan, 2020). Regions in the input image contributing most to the classification result are highlighted in the heat map. From several methods currently available generating heat maps (Ribeiro et al., 2016; Simonyan et al., 2014; Springenberg et al., 2015; Zeiler and Fergus, 2014; Zintgraf et al., 2017), we based our proposed method on the deep Taylor decomposition (DTD) method (Montavon et al., 2017) which is a special case of layer-wise relevance propagation (LRP) (Bach et al., 2015). LRP, has a solid theoretical framework, has been extensively validated (Montavon, 2019; Samek et al., 2017) and can be efficiently implemented, enabling online heat map generation during training.

Besides indications from aforementioned studies, our experiments on Alzheimer’s disease classification showed that CNNs might learn from (misleading) features outside the parenchyma or features introduced by the skull stripping algorithm. Thus, besides investigating how preprocessing steps including registration and skull stripping identify relevant features, we additionally present a novel relevance guided algorithm, mitigating the necessity and impact of skull stripping for classification of brain diseases. Based on its implementation this is referred to as Graz^+^ technique (guided relevance by adaptive *z*^+^-rule).

In summary, the specific contributions of this work are:

- CNN-based disease classification in a cohort of 128 patients with AD and 290 age-matched normal controls.
- Using subject-level 3D T1-based MR image data, differently preprocessed regarding registration and skull stripping.
- Graz^+^ technique: A relevance-guided regularization technique for CNN classifiers to mitigate the impact of MRI preprocessing.
- Making the framework’s source code freely available for reproducibility of the presented results.

## Methods

### Subjects

Inclusion criteria for all participants was a diagnosis of probable or possible AD according to the NINCDS-ADRDA criteria (Knopman et al., 2001) and a complete MRI and study protocol as described in detail in (Damulina et al., 2020). The healthy control (HC) group was selected from participants of a study in community-dwelling individuals. These volunteers were randomly selected from the community register, had a normal neurological status, and were without cerebrovascular attacks and dementia as previously described (Schmidt et al., 2003). This study was approved by the ethics committee of the Medical University of Graz (IRB00002556) and signed written informed consent was obtained from all study participants or their caregivers. The trial protocol for this prospective study was registered at the National Library of Medicine (trial identification number: NCT02752750).

### MR imaging

Patients and controls were scanned using a consistent MRI protocol at 3 Tesla (Magnetom TimTrio; Syngo MR B17; Siemens Healthineers, Erlangen, Germany) using a 12-channel phased-array head coil. Structural imaging included a T1-weighted 3D MPRAGE sequence with 1 mm isotropic resolution (TR/TE/TI/FA = 1900 ms/2.19 ms/900 ms/9°, matrix = 176×224×256) and an axial FLAIR sequence (resolution of 1×1×3mm^3^) for the assessment of white matter abnormalities.

### Data selection

Totally 132 patients with probable AD with 295 scans (Damulina et al., 2020) and 381 controls with 514 scans from an ongoing community dwelling study (Schmidt et al., 2003) were included in this retrospective study. From patients we excluded 12 MRIs because T1-weighted images were not available, and 14 scans because the image matrix was differently sized. Similarly, from controls we excluded 13 MRIs because of missing T1-weighted images, and 17 MRIs because of different image matrix sizes. Age-matching was achieved by excluding 5 scans of patients and 106 scans of controls, yielding 264 T1-weighted images from 128 patients with probable AD (mean age=71.9±8.5 years) and 378 MRIs from 290 healthy controls (mean age=71.3±6.4 years) for the subsequent deep learning analysis.

### Preprocessing

Brain masks from T1-weighted MRIs were obtained using BET from FSL 6.0.3 with bias field/neck cleanup enabled and a fractional intensity threshold of 0.35 (Smith et al., 2004). T1-weighted images were registered to the MNI152 T1 template (**A**) affinely, using FSL flirt with 6 degrees of freedom and a correlation ratio based cost function, and (**B**) nonlinearly, using FSL fnirt with the *T1_2_MNI152_2mm* configuration.

### Attention mask

Our relevance-guided method is preconditioned by binary attention masks. We used entire brain masks obtained by FSL-BET to focus the classifiers to the intracranial volume.

### Classifier network

We based our classifier on the 3D subject-level classifier network in (Wen et al., 2020). Although the proposed network is reported to perform quite well, the number of trainable parameters (42 millions) relative to the dataset size is high, thus rendering it prone to overfitting. Hence, the number and size of the convolutional and fully connected layers were reduced until the network stopped overfitting on the training data and the validation accuracy started to drop. Batch normalization layers did not influence the performance of the network and were therefore removed. Finally, we replaced the max pooling layers by convolutional layers with striding as tested in (Springenberg et al., 2015). Avoiding max pooling layers improves the interpretability of networks (Montavon et al., 2018). Dropout was not applied in the network and all biases were constrained to be negative or zero. The final 3D classifier network is combining a single convolutional layer (kernel 3×3×3, 8 channels) with a down-convolutional layer (kernel 3×3×3, 8 channels, stridding 2×2×2) as the main building block. The overall network stacks 4 of these main building blocks followed by two fully connected layers (16 and 2 units) with totally 0.3 million trainable parameters. Each layer is followed by a Rectified Linear Unit (ReLU) nonlinearity, except for the output layer where a Softmax activation is applied.

### Heat mapping

Heat maps were created based on the deep Taylor decomposition (DTD) method described in (Montavon et al., 2017). This method is equivalent to the layerwise relevance propagation rule LRP-*α*_1_*β*_0_ for networks like the one we used in this study. The principal idea of DTD is to compute a Taylor decomposition of the relevance at a given network layer onto the lower layer. The name “deep Taylor decomposition” comes from the iterative application of Taylor decomposition from the top layer down to the input layer (Montavon et al., 2018). The output of the Softmax layer of the classifier network defines the relevance that is redistributed with this saliency method. With DTD the relevance is routed only along the positively contributing parts of the network. This is a desired property because we want to focus the network on brain regions with features that cause the classification result. Nevertheless, it is of importance to select a heat mapping method that passes simple sanity checks and is dependent on the network and the training (Adebayo et al., 2020; Yona and Greenfeld, 2021). While lower layers could become less influencing on the saliency map (Sixt et al., 2020), DTD was shown to pass the sanity checks by computing saliency maps for all classes and then removing less relevant pixels from the final map (Gupta and Arora, 2019). Due to the nature of brain MRI data, we extended the currently available implementation of DTD from (Alber et al., 2019) to full 3D. The heat mapping method is used for both the relevance-guided classifier network and visualization.

### Relevance-guided classifier network

The proposed relevance-guided network architecture focuses the classifier network on *relevant features* by extending the given network (cf. Figure 1 top) with a relevance map generator (cf. Figure 1 bottom). To this end we implemented the deep Taylor decomposition (*z*^+^-rule) to generate the relevance maps of each input image depending on the classifier’s current parameters during training, yielding the Graz^+^ technique (guided relevance by adaptive *z*^+^-rule).

**Fig. 1.**
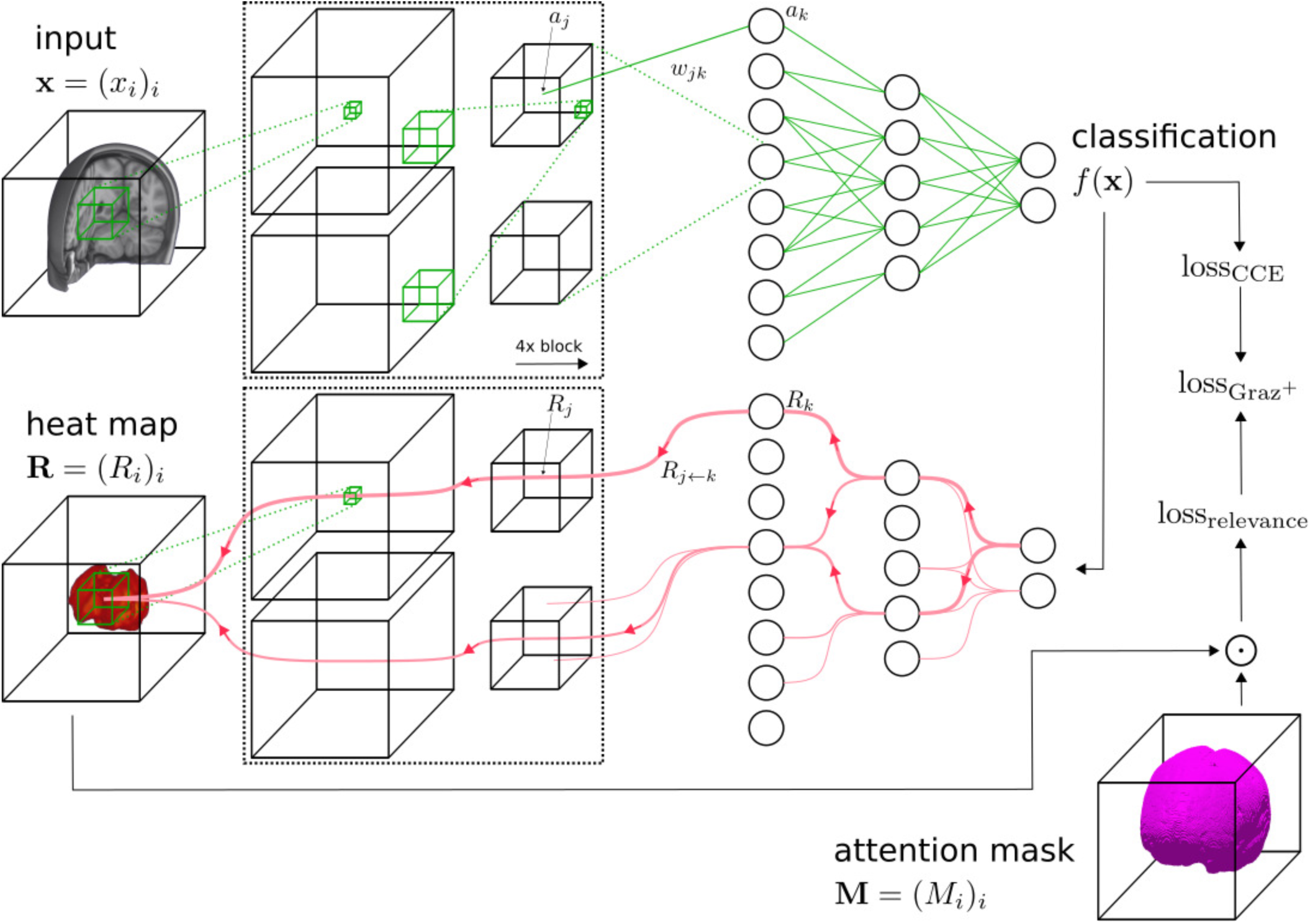
Schematic overview of the Graz ^+^ network and the adapted training process. A conventional classifier network (top) is extended by the heat map generator (bottom). For each classifier network layer a corresponding relevance redistribution layer with shared parameters and activations is attached to the generator network. The online calculated heat map is guiding the classifier training by adding a relevance sum inside the binary attention mask (lossrelevance), which is added to the categorical cross entropy loss (loss_CCE_), yielding the total loss (lossGraz+). *0* denotes the element-wise product.

### Loss function

In order to guide the training process by the attention mask (**M**), we extended the classifier’s categorical cross entropy loss (loss_CCE_) by a relevance-guided loss term to act as a regularizer:

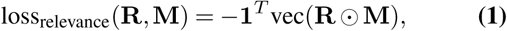

consequently yielding the total loss per data sample:

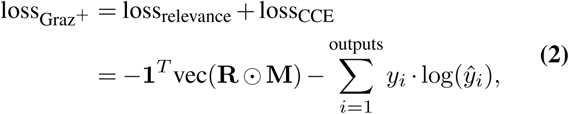

where **R** denotes the relevance heat map (3D shape), **M** is the predefined binary attention mask obtained during image preprocessing (3D shape), vec(**A**) denotes the row major vector representation of **A** resulting in a column vector (1D shape), and **1** is a column vector where all elements are set to 1 (1D shape). The inner product of the transposed vector **1** and the vector representation of **R ⊙ M** gives the scalar value loss_relevance_ (0D shape). The negative sign accounts for the maximization of the relevance inside the mask and **⊙** denotes the element-wise product. For the categorical cross entropy *y*_*i*_ is the target value of the *i*-th output class and *ŷ* _*i*_ its predicted value.

### Hyperparameter optimization and training

Hyperparameter search on learning rate and learning rate schedule was done as proposed in CIFAR10-VGG11 experiments, in detail described in (Bouthillier et al., 2021). The batch size was omitted for consistent memory usage and exponential decay applied for learning rate schedule. Briefly, the hyperparameter optimization resulted in using the Adam optimizer with learning rate set to 10 ^*−*4^, *γ* set to 1.0, *β*_1_ set to 0.9, *β*_2_ set to 0.999 and *E* set to 10 ^*−*7^ (Kingma and Ba, 2015) for 60 epochs with a batch size of 8 for training in all configurations. Each model was end-to-end trained with standard loss minimization and error backpropagation. We trained models for 3 differently preprocessed T1-weighted input images

- in native subject space,
- linearly registered to MNI152 template and
- nonlinearly registered to MNI152 template

and all cases were tested in

- standard classifier network with native images,
- standard classifier network with the skull removed and
- our relevance-guided method with predefined attention masks,

creating overall nine models. No data augmentation was used.

### Cross validation

AD and HC data were split up randomly into five folds without a separate test set, while maintaining all scans from one person in the same fold (Wen et al., 2020). Final folds were created by combining one fold from each cohort to ensure class distribution within. The difference in the class sizes was accounted for using a class weighting in the loss function.

### Model selection

The optimal models based on the standard classifier networks were selected by highest validation classification accuracy. The relevance inside the attention mask threshold was set to 90% for the Graz^+^ networks, enforcing models where most of the relevance is inside the intracranial volume.

### Relevance-weighted heat map representation

Besides qualitatively investigating individual heat maps, we calculated mean heat maps and histogram for each mean heatmap. Starting with the bin with the highest relevance values, the bin contents were added up until 50% of all relevance was included. The lower value of the last bin added was used as the lower value for windowing the mean heatmap. All heat maps shown in this paper are overlaid on the MNI152 1mm template and windowed to present the top 50% of relevance.

### Relevance density

The relevance density describes the contribution of individual voxels of the heat map to the classification result. For all models we compare how many voxels are necessary to reach a certain level of explanation, e.g. how many voxels are needed to explain 85% of the total relevance.

### Volumetry

For comparison between deep learning and logistic regression models for AD classification, we calculated whole brain, gray matter as well as ventricular volume using FSL-SIENAX with a fractional intensity threshold of 0.35 and bias field/neck cleanup enabled (Smith et al., 2002).

### Source code and data availability

Source code for Graz^+^ and the image preprocessing is available under www.neuroimaging.at/explainable-ai. The MR images used in this paper are part of a clinical data set and therefore are not publicly available. Formal data sharing requests will be considered.

## Results

### Model performances

Table 1 reports the mean performance for the cross validation setup of all tested configurations. In summary:

- While models with skull stripping perform better than those without, the Graz^+^ models yield even better balanced accuracy.
- The Graz^+^ model with linearly registered input had the highest balanced accuracy (86.19%), AUC (0.92) and also specificity (92.66%).
- Linear and nonlinear registration improves the balanced accuracy independently of skull stripping and utilization of Graz^+^.
- The logistic regression model based on volumetric information for the entire brain, gray matter, and ventricular volume yielded a balanced accuracy of 82.00%, which is comparable or even outperforming some CNN models without skull stripping.

**Table 1.** Mean performance (in %) for the different models on all holdout data sets of cross validation. Highest values per column are highlighted in bold. *logistic regression by FSL-SIENAX (BET + tissue segmentation) **linear registration is applied during FSL-SIENAX processing to obtain scaling factor AUC, area under the curve of the receiver operating characteristics.

| Classifier | Skull stripping | Registration | Balanced accuracy | Sensitivity | Specificity | AUC |
| --- | --- | --- | --- | --- | --- | --- |
| CNN | no | - | 71.26±2.86% | 55.55±7.51% | 86.96±3.95% | 0.75±0.02 |
|  |  | lin. | 74.27±3.83% | 63.13±9.05% | 85.40±6.45% | 0.80±0.05 |
|  |  | nonlin. | 77.61±4.44% | 64.79±5.02% | 90.43±5.19% | 0.85±0.06 |
| CNN | yes | - | 77.66±4.39% | 69.70±7.65% | 85.63±4.06% | 0.83±0.05 |
|  |  | lin. | 79.45±3.34% | 76.87±4.81% | 82.03±6.23% | 0.86±0.05 |
|  |  | nonlin. | 82.13±5.08% | 73.47±7.89% | 90.78±4.92% | 0.88±0.05 |
| CNN+Graz<sup>+</sup> | no | - | 80.66±4.80% | 74.95±7.85% | 86.36±2.85% | 0.88±0.04 |
|  |  | lin. | **86.19±6.01%** | 79.73±10.72% | **92.66±3.73%** | **0.92±0.04** |
|  |  | nonlin. | 83.50±5.90% | 77.16±8.95% | 89.83±4.49% | 0.90±0.04 |
| Logistic Regression* | yes | lin.** | 82.00±4.25% | **80.57±7.16%** | 83.43±2.45% | 0.90±0.04 |

As the used dataset is nearly balanced (Saito and Rehmsmeier, 2015), the corresponding mean receiver operating characteristics (ROC) curves for these models are shown in Figure 2.

**Fig. 2.**
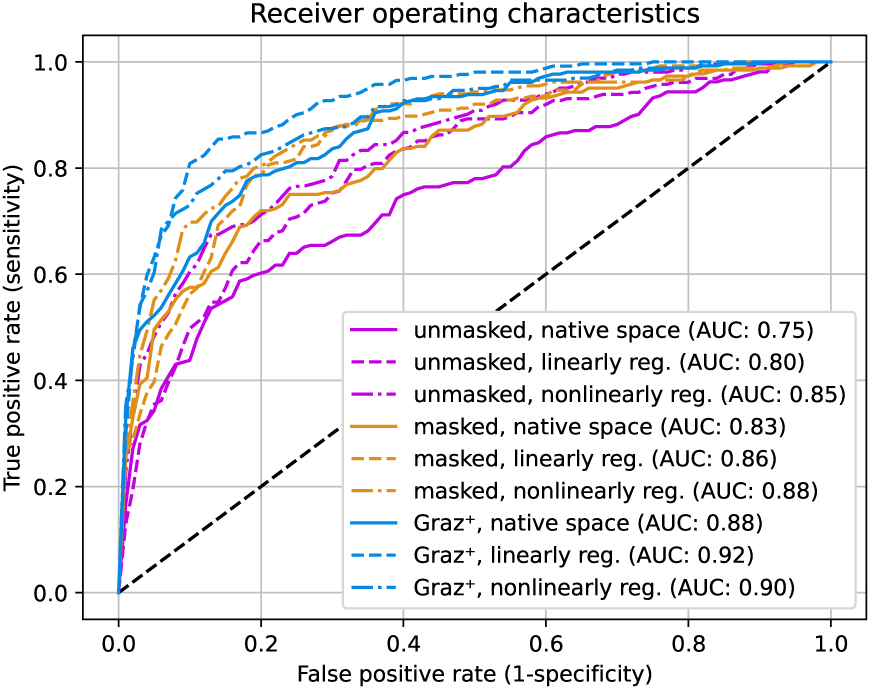
Comparison of mean receiver operating characteristics curves for all nine configurations. The Graz ^+^ models (blue) show higher values for the area under the curve (AUC in legend) compared to unmasked (purple) and masked (orange)

### Heat mapping

Mean heat maps for classification decisions on cross validation holdout data sets for all trained models are shown in Figure 3, overlaid on the MNI152 1mm template. Individual heatmaps were nonlinearly transformed to the MNI152 space before averaging. Transformation information was obtained during T1-weighted image preprocessing. Visual inspection of the heat maps reveals that the processing type (unmasked/masked/Graz^+^) yields substantially different results (columns), while the impact of the registration type (no registration/linear/nonlinear) is rather limited. Although mean heat maps in each column appear visually similar, applying registration to input MRIs improves the balanced accuracy. When using the native T1-weighted images as input, the most relevant features are obtained in the scalp/skull outside brain parenchyma (unmasked configurations, left column). When skull stripping of the input MRIs is applied, the highest relevances are found in the cerebral and cerebellar cortex or generally adjacent to the brain-CSF-interface (middle column). While the aforementioned classifiers also show minor relevances in central brain regions, the maps from Graz^+^ show relevant regions exclusively within deep gray and white matter tissue adjacent to the ventricles (right column). Figure 4 shows multiple slices of mean heat maps for classification decisions of all cross validation holdout data sets for all trained models, overlaid on the MNI152 template.

**Fig. 3.**
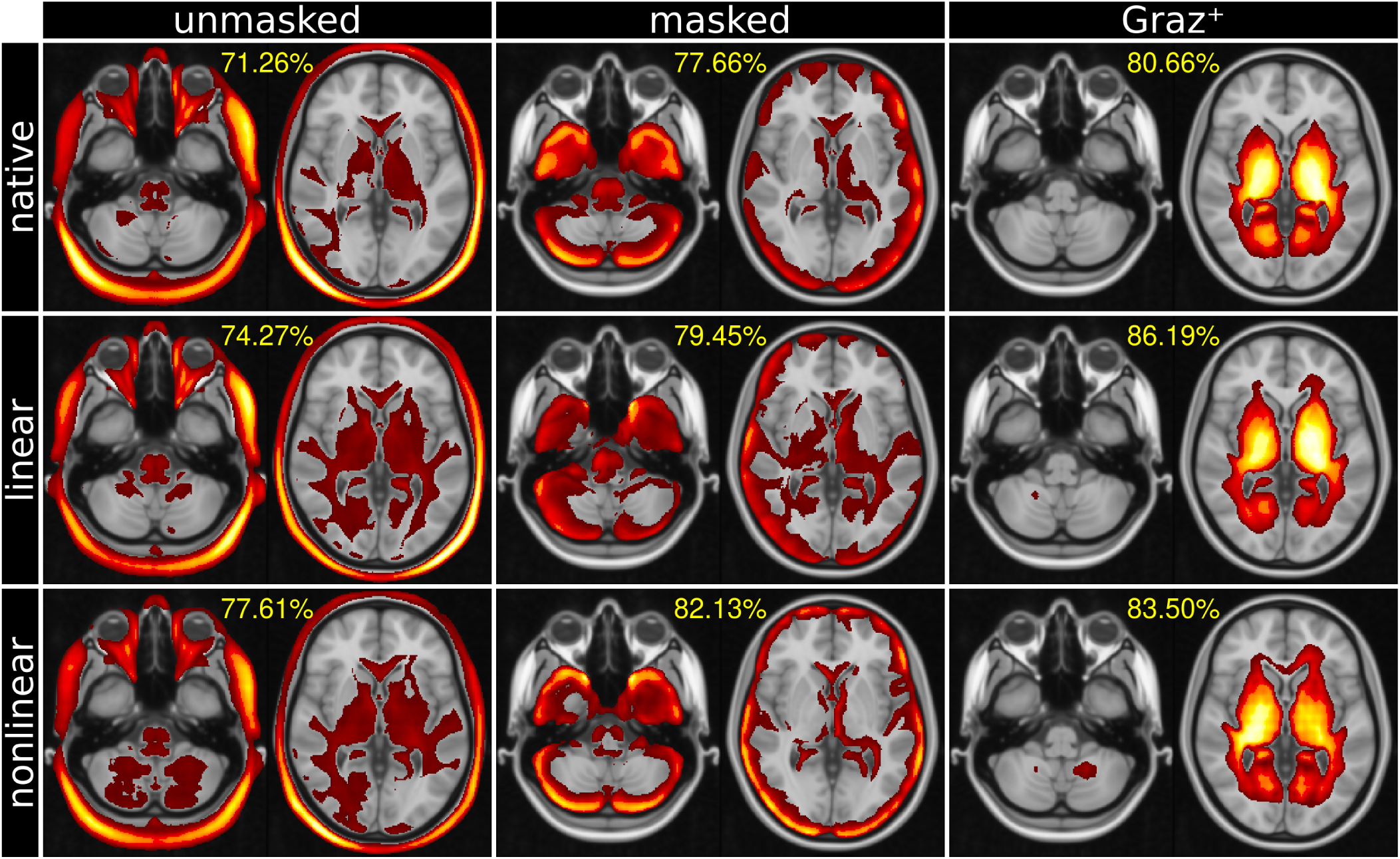
Mean heat maps (highest relevance in yellow, overlaid on MNI152 template) and balanced classification accuracy (percentage). Unmasked and masked CNN classifiers obtain relevant image features overwhelmingly from global volumetric information (left and center columns), whereas Graz ^+^ exclusively relies on deep gray and white matter tissue adjacent to the ventricles (right column). Heat maps are thresholded to the top 50% of the overall relevance. See Methods for description.

**Fig. 4.**
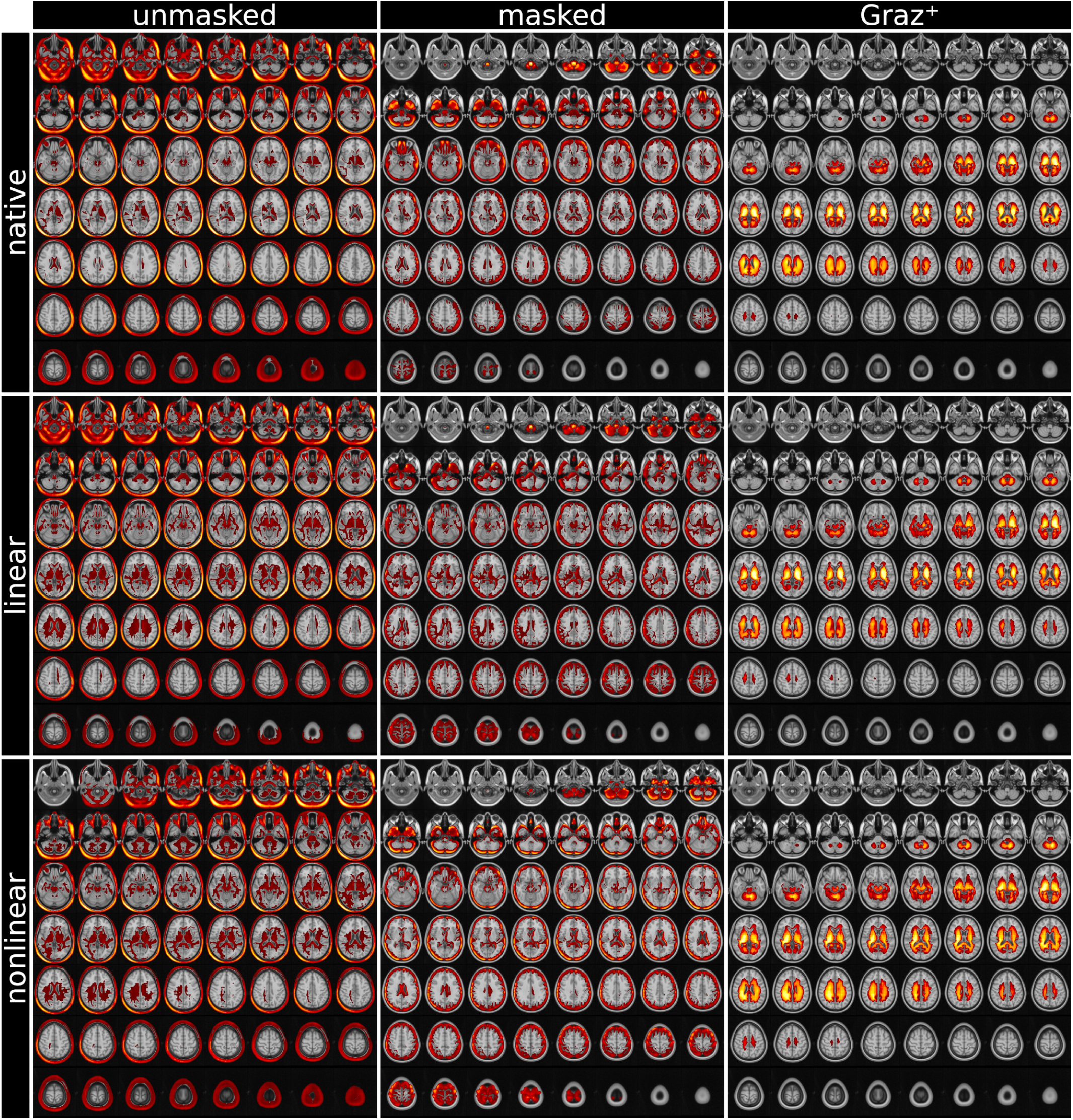
MNI152 template overlaid by mean relevance maps (highest relevance in yellow) obtained for all nine models. Unmasked and masked MRI classifiers obtain relevant image features from volumetric information (left and center columns). In contrast, the proposed Graz ^+^-method bases the classifier’s decision on deep brain image features, virtually independently of the registration method (right column). Heat maps are thresholded to the top 50% of the overall relevance. See Methods for description.

### Relevance density

Figure 5 shows that the Graz^+^ training increased the sparsity of the utilized features, where the 10% most relevant voxels (x-axis) explain approximately 20% (unmasked), 35% (masked) and 75% (Graz ^+0^) of the total relevance.

**Fig. 5.**
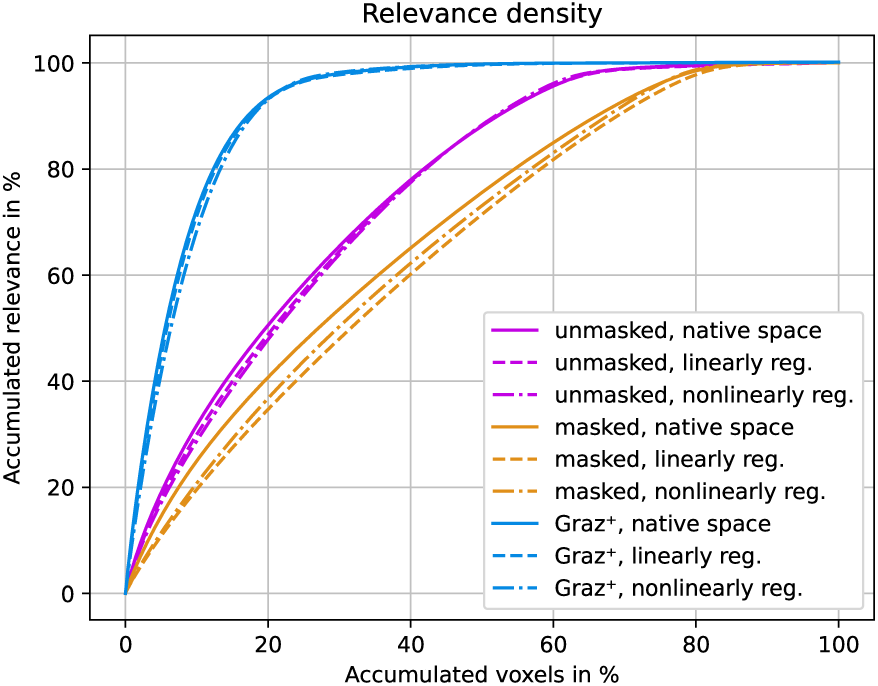
The relevance density describes the contribution of individual voxels to the classification decision. Removal of scalp tissue voxels (orange) yields higher relevance density compared to unmasked T1 images (purple). The Graz ^+^-models (blue) identify sparser but substantially more relevant voxels, which improves the classification accuracy.

## Discussion

### Summary

The present work investigated the mechanisms underlying brain disease classification by CNNs. Understanding the classifier’s decision(s) is highly relevant, not only from an ethno-clinical but particularly from a legal perspective. We demonstrated how dramatic T1-weighted Alzheimer’s disease classification is depending on volumetric features. Moreover, we show that preprocessing of neuroimaging data is decisive for feature identification because it introduces novel misleading features subsequently utilized for classification. The presented Graz^+^ technique is addressing these issues by focusing the feature identification on the intracranial space only. This yields higher classification accuracy than conventional CNN-methods, but more importantly, it substantially resolves the impact of MR image preprocessing.

### Impact to deep learning-based neuroimaging studies

Our motivation for this work was driven by simple recurring questions in clinical brain MRI studies: Should the skull from a conventional T1-weighted MRI be stripped for further processing or should the entire MRI including skull and neck be usedã Additionally, whether and which type of image registration is required or best as the next preprocessing stepã Showing that the preprocessing of MR images is crucial for the feature identification by CNNs has severe implications for neuroimaging based machine learning classifications. A majority of analysis pipelines apply skull stripping during image processing. This avoids the identification of features outside of the brain tissue, but in turn introduces new edges at the newly created brain mask, which might be subsequently used by the CNN for classification. We anticipate that decisions also might be misled by underlying contributors such as the implementation of the skull stripping algorithm, brain atrophy, but also might reflect visually not observable information as involuntary patients’ movements. Generally, the

source and extent of the newly introduced features remains unclear, however it was demonstrated that skull stripping algorithms can be biased by the patient cohort (Fennema-Notestine et al., 2006), thus, additionally biasing the classification.

Addressing these shortcomings, the proposed relevance-guided Graz^+^ method identified regions of highest relevance in brain parenchyma while the balanced accuracy remained comparable or even better. Moreover, pooling data from rare diseases or generally small datasets often yield potentially spurious results and low replicability (Varoquaux, 2018). Its invariance from registration and skull stripping methods provides a usable method for CNN-based classification studies which might be practically useful when pooling data from different scanners and sites (Clarke et al., 2020) or assisting statistical harmonization (Dinsdale et al., 2021b; Pomponio et al., 2020).

### Neuroanatomical and Biophysical Interpretation

This section highlights plausible mechanisms underlying CNN-based disease classification in AD by analyzing the neuroanatomical position of voxel relevance observed by heat mapping. The highest relevances were observed in the scalp for the CNN models using native (unmasked) input images. With skull stripping (masked), the most relevant voxels were found at the brain-CSF-interface, respectively, at the newly-introduced edges of the brain parenchyma. Anatomically, these regions are substantially overlapping with cortical gray matter, where atrophy is a well-known effect in AD. Cortical gray matter changes might be reflected in the masked CNNs decision, but seem rather implausible because of the small magnitude compared to global atrophy and ventricular enlargement. However, we cannot entirely rule out a secondary effect from the brain extraction algorithm biased by the patient cohort (Fennema-Notestine et al., 2006). Both CNN methods also identified some relevant voxel clusters in deep gray and white matter adjacent to the lateral ventricles (center of the brain), which were substantially smaller. Given the spatial distribution of the relevances, we argue that the two conventional CNN models are overwhelmingly sensitive for global volumetric features. Further evidence therefore comes from the complementary volumetric analysis using an established neuroimaging tool for brain segmentation (FSL-SIENAX) in a logistic regression model. The obtained balanced accuracy of 82% is on par with the top CNN results. Here the question arises whether these computational expensive CNNs just resample a refined volumetric measurementã The Graz^+^-based models identified regions with highest relevance mainly in deep gray and white matter located adjacent to the lateral ventricles. However, the anatomical/biophysical underpinnings of the decisions are less clear than in the conventional CNN models. Beside aforementioned contributions of volumetric features (AD progression is commonly paralleled by ventricular enlargement and global atrophy) also the T1-weighted contrast can pathologically change in AD (Besson et al., 1985). White matter hyperintensities (WMH) are commonly seen in brain MRI in older people and beside their underlying heterogeneous histopathology, they represent radiological correlates of cognitive and functional impairment (Prins and Scheltens, 2015). In a previous study, we found WMHs preferentially in a bilateral periventricular location, partly overlapping with the regions identified here by the Graz^+^-based models (Damulina et al., 2019). Furthermore, other plausible contributors are increased brain iron deposition in the deep gray matter (basal ganglia) of AD patients (Damulina et al., 2020) or cumulative gadolinium deposition of macrocyclic contrast agents (Kanda et al., 2014). Nevertheless, with the given setup we cannot definitely disentangle the underlying constituents and refer to the validation section below.

The relevance density analysis revealed that Graz^+^-based models learn much sparser features, subsequently needing less voxels for inferring classification decisions. Consequently, we hypothesize that the lack of misleading voxels from the scalp or newly-introduced edges is responsible for the increased accuracy.

### Related work

With the availability of accessible large MRI databases from patients, such as the Alzheimer’s Disease Neuroimaging Initiative (ADNI), AIBL or OASIS databases, various studies using machine learning techniques exploiting structural imaging data have been published, formerly using *classical* machine learning classification methods (e.g. LDA, SVM) in combination with feature extraction methods based on tissue density (Klöppel et al., 2008), cortical surface (Eskildsen et al., 2013) and hippocampal measurements (Sørensen et al., 2016). Reported classification accuracies range between 75% and 100%, comprehensively summarized in (Rathore et al., 2017). Recently, interests switched to deep learning CNNs for (**A**) classification (Bäckström et al., 2018; Noor et al., 2020; Zhang et al., 2020), (**B**) classification with explanation (Böhle et al., 2019; Tang et al., 2019; Oh et al., 2019) and (**C**) regression with explanation (Dinsdale et al., 2021a) of AD. A recent review summarizes the state-of-the-art using CNNs for AD classification, comparing various network architectures, input data and disease subtypes (Wen et al., 2020). Strictly in line with the data leakage analysis in this work we utilized stratified cross validation, while maintaining all datasets from one person in the same fold. Furthermore, we used the input MR images in their native spatial resolution, avoiding unpredictable influence from down- or resampling. While most of the analyzed studies are based on the ADNI dataset, our classification performance results are on par with both remaining 3D subject-level approaches without data leakage (Bäckström et al., 2018; Korolev et al., 2017).

The inconsistency between learned features with linear and nonlinear registration is systematically investigated in (Dinsdale et al., 2021a). They found that the use of nonlinearly registered images to train CNNs can drive the network by registration artifacts. However, the influence of further preprocessing steps on the resulting models and performances is less well known. Heat mapping using the LRP framework has been sparsely applied for explaining the underpinnings of an AD diagnosis in convolutional neural networks trained with structural MRI data beside the work of (Böhle et al., 2019). Heat maps obtained by two techniques (LRP and guided backpropagation) indicate relevant features in the parahippocampal gyrus but also adjacent to the brain-CSF interface, which is in line with our work.

Regularized heat map learning has been proposed before, however, differently to the Graz^+^ method integrating a-priori knowledge with predefined attention masks. Technically, the gradient of the function learned by the network with respect to the current input can be interpreted as a heat map (Simonyan et al., 2014). Regularization of this input gradient was first introduced by (Drucker and Le Cun, 1992) as *double back-propagation*, which trains neural networks by not only minimizing the *energy* of the network but the rate of change of that energy with respect to the input features. In (Ross et al., 2017) this regularization was extended by selectively penalizing the gradient. Whereas (Sun et al., 2021) use LRP to create maps during training, which are multiplied with the corresponding input and then fed to the original classifier to dynamically find and emphasize important features. Furthermore, attention gated networks for medical image analysis have been proposed to automatically learn to focus on target structures of varying shapes and sizes (Schlemper et al., 2019).

### Validation

Direct validation of the classifier’s decision is generally hardly feasible in the absence of a ground truth. While we anticipate a correspondence of the volumetric features with Alzheimer’s atrophy, this conclusion might not be final. However, in future work, indirect validation is possible using quantitative MRI parameters such as relaxometry, susceptibility, or magnetization transfer, where regional effects are known from ROI-based, voxel-based morphometry (VBM) or radiomics studies. While those methods statistically assess neuroanatomical features including ventricular enlargement or hippocampal atrophy, quantitative MRI parameters describe the underlying biophysical tissue composition. The effective relaxation rate 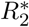 can assess increased iron deposition in the basal ganglia, a frequent finding in AD (Damulina et al., 2020). Consequently, the potential overlap with heat maps in those regions is better suited to disentangle biophysical tissue changes from atrophy. Optionally, direct validation of our method would require the generation of a cohort of realistic in silico phantoms (as recently used in the quantitative susceptibility mapping (QSM) image reconstruction challenge 2.0 (Marques et al., 2021)) with modulateable regional relaxation times in conjunction with an adjustable atrophy deformator (Khanal et al., 2017, 2016).

### Limitations

Several aforementioned neuroimaging studies used the ADNI (or other publicly available) database for deep learning based classification. Generally, the clinical relevance of an automated AD classification is limited. The prodromal state of mild cognitive impairment (MCI) is preceding AD and identification of individuals rapidly progressing to AD (or differential diagnosis of frontotemporal dementia types) would be of higher importance for clinical management. We acknowledge the absence of an MCI group as a limitation and therefore provide the source code for the fast reproducibility using alternative network topologies, input data (quantitative MRI, PET), and other diseases. While aforementioned databases are designed multi-centrically, all MRI scans used in this paper were acquired with a single 3T scanner. Beside the underlying AD patient data, comparison with other studies is hampered by different network architectures, preprocessing and hyperparameter selection (Wen et al., 2020).

While this study only applied whole brain masks, more focused masks guiding the attention to e.g. the precuneus, the entorhinal cortex, the parietal lobe, the temporal lobe or the hippocampi are feasible, especially when regional a-priori knowledge for a certain pathology exists. Because of the explorative nature of the novel methodological framework we focused on the brain parenchyma. Organs outside the brain are more variable in size and shape, which render registration and ROI-definition more challenging. We originally developed Graz^+^ for clinical brain studies, but its invariance to preprocessing might be even more pronounced beyond neuroimaging.

Lastly, the absence of CSF biomarkers or amyloid/Tau-PET for the AD diagnosis reduces the accuracy of the clinical diagnosis. However, AD diagnosis using the NINCDS-ADRDA criteria has a sensitivity of 81% and specificity of 70% as shown in clinico-pathological studies (Knopman et al., 2001).

## Conclusion

This work highlights that CNNs are not necessarily more efficient or better regarding classification accuracy than simple conventional volumetric features. However, the proposed relevance-guided approach is neutralizing the impact of MRI preprocessing from skull stripping and registration, rendering it a practically usable and robust method for CNN-based neuroimaging classification studies. Relevance-guiding focuses feature identification on the intracranial space only, yielding physiological plausible results and as a secondary effect the classification accuracy is higher.

## Data Availability

Source code for Graz+ and the image preprocessing is available under www.neuroimaging.at/explainable-ai. The MR images used in this paper are part of a clinical data set and therefore are not publicly available. Formal data sharing requests will be considered.

## ACKNOWLEDGEMENTS

This study was funded by the Austrian Science Fund (FWF grant numbers: KLI523, P30134). This research was supported by NVIDIA GPU hardware grants.

## Appendix A

Table A.1 shows search space and default values for the hyperparameter optimizations for all configurations.

## Appendix B

Table B.1 shows performance for the different models on all holdout data sets of cross validation.

**Table A.1.** Search space and default values for the hyperparameter optimizations for all configurations.

| Hyperparameter | Default | Space |
| --- | --- | --- |
| learning rate | $10^{-4}$ | $\log(10^{-5}, 10^{-3})$ |
| $\gamma$ of learning rate schedule | 0.997 | $\text{lin}(0.96, 1.0)$ |
| $\beta_1$ | 0.9 | - |
| $\beta_2$ | 0.999 | - |
| batch size | 8 | - |

**Table B.1.** Performance (in %) for the different models on all holdout data sets of cross validation. *logistic regression by FSL-SIENAX (BET + tissue segmentation) **linear registration is applied during FSL-SIENAX processing to obtain scaling factor AUC, area under the curve of the receiver operating characteristics. 14 Christian Tinauer *et al*. | Graz ^+^

| Classifier | Skull stripping | Registration | Cross validation index | Balanced accuracy | Sensitivity | Specificity | AUC |
| --- | --- | --- | --- | --- | --- | --- | --- |
| CNN | no | - | 1 | 73.28% | 54.90% | 91.67% | 0.74 |
|  |  |  | 2 | 67.02% | 48.15% | 85.90% | 0.72 |
|  |  |  | 3 | 70.19% | 49.02% | 91.36% | 0.77 |
|  |  |  | 4 | 70.41% | 56.60% | 84.21% | 0.78 |
|  |  |  | 5 | 75.39% | 69.09% | 81.69% | 0.76 |
| CNN | no | lin. | 1 | 77.49% | 64.71% | 90.28% | 0.76 |
|  |  |  | 2 | 67.81% | 57.41% | 78.21% | 0.75 |
|  |  |  | 3 | 73.24% | 58.82% | 87.65% | 0.84 |
|  |  |  | 4 | 74.07% | 54.72% | 93.42% | 0.80 |
|  |  |  | 5 | 78.73% | 80.00% | 77.46% | 0.87 |
| CNN | no | nonlin. | 1 | 83.62% | 68.63% | 98.61% | 0.92 |
|  |  |  | 2 | 71.94% | 59.26% | 84.62% | 0.78 |
|  |  |  | 3 | 75.93% | 66.67% | 85.19% | 0.80 |
|  |  |  | 4 | 74.64% | 58.49% | 90.79% | 0.83 |
|  |  |  | 5 | 81.93% | 70.91% | 92.96% | 0.91 |
| CNN | yes | - | 1 | 80.15% | 68.63% | 91.67% | 0.84 |
|  |  |  | 2 | 70.66% | 59.26% | 82.05% | 0.76 |
|  |  |  | 3 | 78.36% | 76.47% | 80.25% | 0.82 |
|  |  |  | 4 | 75.50% | 64.15% | 86.84% | 0.80 |
|  |  |  | 5 | 83.66% | 80.00% | 87.32% | 0.90 |
| CNN | yes | lin. | 1 | 78.35% | 70.59% | 86.11% | 0.89 |
|  |  |  | 2 | 77.14% | 72.22% | 82.05% | 0.80 |
|  |  |  | 3 | 76.98% | 82.35% | 71.60% | 0.81 |
|  |  |  | 4 | 78.81% | 77.36% | 80.26% | 0.86 |
|  |  |  | 5 | 85.98% | 81.82% | 90.14% | 0.93 |
| CNN | yes | nonlin. | 1 | 89.09% | 82.35% | 95.83% | 0.93 |
|  |  |  | 2 | 74.07% | 64.81% | 83.33% | 0.79 |
|  |  |  | 3 | 79.99% | 68.63% | 91.36% | 0.85 |
|  |  |  | 4 | 81.99% | 67.92% | 96.05% | 0.93 |
|  |  |  | 5 | 85.48% | 83.64% | 87.32% | 0.90 |
| CNN+Graz<sup>+</sup> | no | - | 1 | 86.32% | 82.35% | 90.28% | 0.92 |
|  |  |  | 2 | 79.06% | 72.22% | 85.90% | 0.88 |
|  |  |  | 3 | 76.03% | 70.59% | 81.48% | 0.81 |
|  |  |  | 4 | 75.50% | 64.15% | 86.84% | 0.84 |
|  |  |  | 5 | 86.39% | 85.45% | 87.32% | 0.93 |
| CNN+Graz<sup>+</sup> | no | lin. | 1 | 92.44% | 86.27% | 98.61% | 0.96 |
|  |  |  | 2 | 89.67% | 87.04% | 92.31% | 0.94 |
|  |  |  | 3 | 75.82% | 62.75% | 88.89% | 0.85 |
|  |  |  | 4 | 83.22% | 71.70% | 94.74% | 0.92 |
|  |  |  | 5 | 89.82% | 90.91% | 88.73% | 0.94 |
| CNN+Graz<sup>+</sup> | no | nonlin. | 1 | 89.79% | 82.35% | 97.22% | 0.93 |
|  |  |  | 2 | 79.13% | 68.52% | 89.74% | 0.89 |
|  |  |  | 3 | 76.54% | 66.67% | 86.42% | 0.84 |
|  |  |  | 4 | 80.78% | 77.36% | 84.21% | 0.88 |
|  |  |  | 5 | 91.23% | 90.91% | 91.55% | 0.94 |
| Log. Regr.* | yes | lin.** | 1 | 80.31% | 74.51% | 86.11% | 0.89 |
|  |  |  | 2 | 76.50% | 72.22% | 80.77% | 0.86 |
|  |  |  | 3 | 79.34% | 78.43% | 80.25% | 0.85 |
|  |  |  | 4 | 86.16% | 86.79% | 85.53% | 0.94 |
|  |  |  | 5 | 87.71% | 90.91% | 84.51% | 0.95 |

